# Beyond Common Outcomes: Client’s Perspectives on the Benefits of Prenatal Care Coordination

**DOI:** 10.1101/2022.04.12.22273331

**Authors:** Madelyne Z. Greene, Bikki Tran-Smith, Pahder Moua

## Abstract

Prenatal Care Coordination (PNCC) is a fee-for-service Medicaid benefit available in Wisconsin and several other states. It provides for home visiting, health education, care coordination and other supportive services to high-risk mothers. However, PNCC is not supported by an evidence-based model, its impact is not systematically evaluated, and the benefit is not currently reaching all eligible mothers. The purpose of this qualitative descriptive study was to describe PNCC clients’ perspectives on the experience of receiving the PNCC benefit and the value and impact of PNCC in the context of their own lives. We interviewed recent clients of PNCC programs in two PNCC sites that varied by racial/ethnic community makeup, rural/urban geography, and health department size. PNCC clients identified two major benefits of PNCC: 1) social and emotional support from the PNCC nurse; and 2) assistance with obtaining or getting connected to other resources they needed. These two program benefits were especially meaningful to PNCC clients in the context of difficult life events and circumstances. Findings from this study highlight the impact of PNCC services on social and emotional health through trusting and supportive nurse-client relationships. Our findings also suggest that a longer program period and the development of standards to assess program effectiveness would improve PNCC client outcomes and reduce disparities in maternal health.

## Introduction

Maternal mortality and severe morbidity have been on the rise in the US over the past several decades (Callaghan, 2020; Petersen, Davis, Goodman, Cox, Mayes, et al., 2019; Petersen, Davis, Goodman, Cox, Syverson, et al., 2019). A complex set of social and structural risk factors—such as poverty, unsafe and inconsistent housing, and exposure to racism— contribute to poor maternal health outcomes. Housing insecurity among pregnant women, for example, which is especially common in minoritized populations such as Black mothers (Leifheit et al., 2020), has been associated with low infant birth weight, poor health outcomes, and extended hospitalization. Strong social support systems, however, can mitigate these risks (Bedaso, Adams, Peng, & Sibbritt, 2021).

As standard medical prenatal care (which itself is inaccessible for many mothers) does not attend to these complex social and structural risk factors, a number of care coordination programs and policies, such as the Medicaid-covered Prenatal Care Coordination (PNCC) benefit have been established to address the gaps in care. This fee-for-service benefit has been created to support access to medical, social, educational, and other pregnancy-related services, and ultimately improve birth outcomes. Several states including Wisconsin, which has the starkest racial disparities between Black and white mothers of any state, offer the PNCC benefit.

The PNCC benefit has been shown to reduce the likelihood of prematurity and low birthweight in Wisconsin (Mallinson, Larson, Berger, Grodsky, & Ehrenthal, 2020; Van Dijk, Anderko, & Stetzer, 2011) and other states (Meghea, You, Raffo, Leach, & Roman, 2015; Ricketts, Murray, & Schwalberg, 2005; Skovholt et al., 1994). However, though PNCC has the potential to improve several infant health outcomes, there is limited research on the effectiveness or outcomes of PNCC on *mothers*. One such study highlighted the importance of the nurse-client relationship and the social support available through this relationship in PNCC on maternal well-being (Heitzman, Weitzel, Kroll, & Zabler, 2019).

There is more evidence on the effectiveness of a longstanding and comparable integrated care model, the Nurse-Family Partnership (NFP). NFP is funded by federal Maternal, Infant, and Early Childhood Home Visiting (MIECHV) grant funds and provides services for first-time mothers from the prenatal period through the infant’s first two years of life.in improving both infant and maternal health outcomes. It has been shown to reduce neonatal mortality, preterm birth, and childhood maltreatment (Miller, 2015; Thorland & Currie, 2017), as well as reduce pregnancy complications and substance use during pregnancy (Olds, 2006; Olds et al., 2019). A strong contributor to these outcomes has been the development of a positive therapeutic nurse-patient relationship enabling nurses to support mothers socially and emotionally and work together with them to meet their specific goals and needs (Landy, Jack, Wahoush, Sheehan, & Macmillan, 2012).

### PNCC Benefit Structure and Processes

Pregnant women are referred to PNCC service providers from a variety of sources, but in Wisconsin, referral commonly occurs through prenatal care providers and the Special Supplemental Nutrition Program for Women, Infants, and Children (WIC) offices. In order to receive services through the PNCC benefit, pregnant people must meet minimum criteria for risk according to the program’s Pregnancy Questionnaire (ForwardHealth, 2009). Criteria include low income, prior history of preterm birth, homelessness or unstable housing, and young maternal age. In contrast to NFP and other more structured programs, clients are eligible to receive PNCC services any time during their pregnancy and for all pregnancies, not just the first. Once screened and enrolled, PNCC providers–Registered Nurses, community health workers, or other care providers–render covered services and then bill the state’s Medicaid program. PNCC services fall under four major categories: assessment, care plan development, care coordination and monitoring, and health education. The benefit’s stated targets are facilitating early initiation of prenatal care, referrals to resources, and providing individual psychosocial support. Beyond these targets there are limited service provision guidelines, affording PNCC providers the flexibility to deliver individualized care and services to meet each client’s needs.

### Study Aims

PNCC, as a fee-for-service insurance benefit, is not supported by a well-developed program or model, and is not systematically evaluated. Additionally, the benefit is not currently reaching all eligible mothers; in a recent study, only 25% of women with Medicaid were evaluated for PNCC benefits (Larson, Berger, Mallinson, Grodsky, & Ehrenthal, 2019). There has also been significant variation in screening and uptake of PNCC services depending on maternal county of residence (Larson et al., 2019). As awareness of the impact of social and structural risk factors on maternal health increases, and additional investments are made in integrated supportive care services, more research is needed to understand PNCC’s impact. Thus, the purpose of this qualitative descriptive study was to describe PNCC clients’ perspectives on the experience of receiving the PNCC benefit and the value and impact of PNCC in the context of their own lives.

## Methods

The overarching study aim was to examine the implementation of PNCC at two comparison program sites—one urban and one rural—in Wisconsin. To achieve this aim, we conducted semi-structured interviews with recent PNCC clients from each site. We also employed ethnographic observation and individual interviews with PNCC providers who were Registered Nurses (RNs) (although not all PNCC providers in the state are RNs) and managerial and support staff at both sites. Here, we report findings from the PNCC clients about their perspectives on the value and implementation of PNCC. This study was approved by the UW-Madison IRB.

### Sites and Sample

Two PNCC programs were selected as study sites for their differences in the size and resource availability of the PNCC agency, urban or rural county, the socio-structurally vulnerable groups that made up the site’s client population, and rates of PNCC eligibility screening and service delivery in the county (Larson et al., 2019). Participation from PNCC providers was optional and nurses had in writing from supervisors that they were not required to participate. However, staff buy-in to the research was high and all nurses participated (five at one site and one at the other site) and expressed enthusiasm for the value of the research.

We recruited former PNCC clients from each site who had completed the program within the previous year. Sampling strategies for clients were unique to each site and informed by the typical communication methods that nurses used with clients, since the trusting relationships built between nurses and community members were identified as critical to the programs. At the larger, urban site, we used a tiered outreach strategy, which included mailing physical letters to all clients who had completed the program within the previous year and following up with text messages from their former PNCC provider. PNCC providers referred clients who were interested in the study to the PI to answer questions and complete the screening process. At the smaller, rural site, PNCC team members individually reached out to the relatively few clients who had completed the program in the previous year, and again referred any interested clients to the research team.

### Data Collection

The data collection process began with observational data collection at each site; the first author observed PNCC nurses, administrative staff, and managers conducting PNCC work, including weekly team meetings, scheduling tasks, billing, planning for and conducting client visits, and documenting visits in patients’ charts. Semi-structured interviews were conducted with each former PNCC client in the months surrounding those observations. Interviews with PNCC clients were offered in-person or via web-based conference per participant preference. All interviews were audio-recorded and transcribed *verbatim*. Each participant received a $30 gift card for the store of their choice for their participation.

### Data Analysis

Inductive thematic analysis was used to code the interview transcripts. (Braun & Clarke, 2006) All three authors did multiple re-readings of the interview transcripts and then conducted line-by-line coding of one interview. Initial coding was compared and discussed to establish a draft codebook. All three authors then coded an additional two transcripts, further refining the codebook and establishing consistency in applying the codes. Two coders then independently coded each subsequent transcript and discussed coding decisions until reaching 100% agreement. Coding discrepancies that could not be resolved were then discussed by the entire three-person study team until consensus was reached. The codebook was continually refined and edited throughout the coding process. Coding was completed using Dedoose version 9.0.17, a web application for managing and analyzing qualitative data (“Dedoose,” 2021).

To enhance methodological rigor and to establish credibility and trustworthiness, we evaluated the data for thematic saturation, conducted multiple re-readings of each interview transcript, triangulated our interpretation of findings, and used an audit trail (Lincoln & Guba, 1985). The broader study’s data collection process included observation and interviews with the PNCC providers and staff members, allowing the investigator to approach client interviews with deep understanding of PNCC from a programmatic perspective. As each interview was completed, the PI noted major themes and topics that had been mentioned and concluded data collection at each site after two interviews that did not introduce any new topics. Triangulation was accomplished through consensus coding and a theme generating process that involved all three authors. We maintained an audit trail documenting all analytical decisions made by the team throughout the data analysis process to ensure that the analysis was dependable (Lincoln & Guba, 1985).

### Positionality Statement

The study team was made up of an assistant professor (PI), a postdoctoral trainee (2^nd^ author), and one undergraduate research assistant (3^rd^ author), along with the partnership of community stakeholders including the state and local leaders of PNCC programs in Wisconsin. The PI and postdoctoral trainee each have several years of experience collecting and analyzing qualitative data regarding sexual and reproductive health experiences from vulnerable and marginalized populations, and the research assistant is a fourth-year nursing student and Certified Nursing Assistant with clinical experience in pediatrics. All authors are women and have experienced a range of needs along the reproductive and maternal health care spectrum. The PI, who conducted all data collection, occupies a position of relative privilege compared to most PNCC clients; she is white, cisgender, urban-dwelling, highly educated, and associated with state’s flagship university. The second and third authors are Asian-American cis-gender women who are also urban dwelling, highly educated, and associated with the state’s flagship university. The entire study team used a health equity framework, which asserts that current social structures such as the healthcare system can exacerbate inequality and injustice in healthcare access and outcomes for vulnerable populations. Thus, to achieve health equity, interventions should go beyond individual level change to target systemic barriers to high-quality care.

## Results

Participants reported overall positive experiences in the PNCC program, which in many cases was unique compared to their other encounters in healthcare or other social service programs. PNCC clients identified two major benefits of program participation, or outcomes of the program. These outcomes were 1) feeling socially and emotionally supported by the PNCC nurse; and 2) getting assistance with obtaining or getting connected to other resources they needed. This was the case for clients from both sites. These two program benefits were especially meaningful to PNCC clients in the context of difficult life events and circumstances. In the sections below, we report PNCC clients’ thoughts about a) the meaning of each outcome, and b) how nurses operationalized and worked toward each outcome. Finally, we report clients’ recommendations for improving or enhancing these program benefits.

**Table 1.** Themes generated by PNCC Client interviews related to PNCC program outcomes.

|  |  |
| --- | --- |
| Theme 1: Receiving Social and Emotional Support |  |
|  | a) The Meaning of Social and Emotional Support to Clients |
|  | Filling in Gaps Left by Difficult Life Events |
|  | b) How PNCC Nurses Operationalized Social and Emotional Support |
|  | Establishing Therapeutic Relationships Through Reliable Presence |
|  | Providing Holistic Care: Knowing the Community |
| Theme 2: Connections to Needed Resources |  |
|  | a) The Meaning of Connections to Needed Resources for Clients |
|  | Resources in the Context of Social and Emotional Support |
|  | b) How Nurses Operationalized Connecting Clients to Needed Resources |
|  | Individualized Care: Flexible Goals and Focus |
| Theme 3: Client's Recommendations for the Program |  |
|  | a) Lengthen Time Frame of PNCC Program |
|  | b) Improve Marketing and Community Knowledge of PNCC |

### Theme 1: Receiving Social and Emotional Support

#### a) The Meaning of Social and Emotional Support to Clients

Having support was overwhelmingly mentioned as a crucial component of program participation. When asked to describe what she perceived as the purpose of the PNCC program as well as what she gained from it, one participant stated that the “the purpose is the support…to make you feel supported” (1-2-9). Another participant reported that having a PNCC nurse to talk to helped allay first-time pregnancy fears.

> It was like, my first time being pregnant, so like, she made me feel comfortable about…because I was like, scared, you know, to go through this pregnancy… She was just there like to, like, someone to talk to, and […] talk to me… like, made me feel comfortable through my pregnancy. (3-2-8)

Another participant explicitly shared how the need for support went beyond assistance for health and other pregnancy-related concerns to more broadly encompass social and emotional support.

> She [PNCC nurse] helps with, you know, checking the weight, making sure like, the development is going properly and stuff like that, and make sure your health is good, not even just your physical health but your mental health, just like somebody to talk with and communicate with, having like, a friend. Not a lot of people have that, so it was good to have someone to talk about. (1-2-6)

For many participants, their existing social and emotional support was either limited or interrupted due to incarceration, discord, illness, or time constraints. For one participant, support from the PNCC nurse meant she felt less alone during her pregnancy, as her husband was away, and she also had an infant at home.

> I was pretty much like a single mom the past ten months. And these guys up here, they were really helpful throughout that. I feel like if I didn’t have their support, I probably would have given up, because I had a hard time. (3-2-1)

Another participant described how the support of her PNCC nurse helped to alleviate the significant stress she was experiencing from a contentious relationship with her baby’s father and a new medical diagnosis:

> …[I got sick] while I was at work, and I woke up in the hospital and they’re like, “Hey. [you’re sick], but you’re pregnant.” I’m like, “Wait. What?” [laughs] I be like— I wasn’t in a relationship…with my child’s father at the time, so I had to tell him…Like, the rest of my pregnancy was very, like, very, very, very stressful and I was already high risk on top of it, arguing with his dad and stuff…And so, [my nurse] was just kinda like an outlet… [she] wasn’t more, like, a “program lady.” She was more, like, someone I can communicate to and somebody who [will] actually, like, help me focus on what I need to do, what’s best for my health, for my child. (1-2-8)

Some participants, including the client quoted above, characterized the relationships they developed with their PNCC nurse as friendships. One participant went even further to describe the PNCC nurse as akin to family:

> Like, even my own family doesn’t understand it. [The PNCC program staff] understand more than like, even my mom or anybody would, so… [My] mom was like, “oh, I had it worse than you.” But that’s not the point. It’s the point that I have a kid, and it’s hard… They’re [PNCC program staff] like—yeah, they’re like, more of my family than most of my family, because all my family is down in [another city]. I moved here like, seven years ago, and now I have basically nobody except for them and my man and my mom… But she’s really sick, too; she’s been sick since I can remember. (3-2-5)

This participant’s parent had an illness that prevented her from offering consistent support. Similarly, another participant reported that although her parent could be helpful, her main support system came from the PNCC program:

> I mean, I had my mom. She was probably … the other person. But she couldn’t help as much. I feel like my main support system came from that program, at least on people to talk to. Like if I even didn’t need anything—I just needed somebody to talk to—they were always there to talk to. (3-2-4)

Even for those who had an existing social support network, the support from a PNCC nurse was a welcomed addition that helped participants not feel like a burden to their family or healthcare providers.

> Just having that extra support that I might not have had with too many of my family or the midwives. Don’t want to bother them all the time. So, having her even coming to my home, where I’m comfortable, and talking about things…it’s pretty comforting and helpful that way. (1-2-10)

Clients often needed a reliable listening ear without feeling like they were a burden.

#### b) How PNCC Nurses Operationalized Social and Emotional Support

Participants stressed that their nurse’s particular attributes, behaviors, and attitudes were critical to receiving effective and meaningful socio-emotional support during their time as PNCC clients. The factors that made clients feel supported were not described as nursing skills *per se* but can be understood in terms of the core nursing skills and functions of establishing therapeutic nurse-patient relationships and providing holistic care.

##### Establishing Therapeutic Relationships Through Reliable Presence

According to participants, the key to nurses building trusting and mutually respectful relationships, and therefore to providing meaningful support, was their reliability. Reliability to participants meant that the nurse followed through on promised visits and tasks and kept track of what the client needed. Only one PNCC client reported a significantly negative experience before switching to a different nurse, and her major concern was a lack of follow through with the first nurse.

> I would tell her what I need, and she won’t write it down or nothing […] I can’t remember, I needed something while I was pregnant, and I asked her for it. And the next visit, she forgot completely about it. She forgot *completely* about it (1-2-5).

In contrast, this same client described how the second nurse demonstrated that she was reliable and could be counted on:

> [She] would let me talk […] and when stuff—when I couldn’t remember what I had to ask she would ask me questions and it would bring it back to my mind. See, that’s why I like [her], because it’s like she knows… and she’ll write it down, and whatever she wrote down—for the next visit, she brought it, and we always talked about new goals. (1-2-5)

Another client described the nurse’s consistent follow-through:

> [She] made sure that everything that we put goals on, from the first time we met, made sure everything was met before she was like, “okay, I think you got this.” (1-2-6)

Nurses also demonstrated their reliability by responding quickly when a client reached out or had a need, even when the request came outside of regular business hours.

> I was able to hit her up and she [would] communicate right back instead of just being a program, where you have nine-to-five hours, and you have to wait for the next day for the person to get back to you, and if they’re busy, they can’t get back to you. (1-2-8)

For some, reliability was strongly linked to the nurse’s consistent and steady presence in the client’s life. As one client put it, the PNCC nurse was “on top of it. She was there. She helped me out a lot.” (1-2-5). Another client described how, though she was wary of the program at first, the nurse’s consistency helped to develop trust between them:

> Well, at first, I don’t know. I was just like, coming here for prenatal visits and she was helping more like, with my rides and throughout my pregnancy. And, I don’t know, our bond just got stronger and stronger. And like, I knew she was someone I could, you know, come to and [get] support […] I think we even like, became friends. And so, yeah… our bond, I don’t know, like, grew. (3-2-8)

As with the overall experience of social and emotional support, for some participants being able to rely on their PNCC nurse was a unique attribute compared to others in their lives or other healthcare and social service providers.

> Not a lot of people want to help you here, but there’s a select few people here. Like at the clinic I don’t get help… they just send me to another person, send me to another person. Like, I’ll call and set up an appointment; they’ll never call me back. But here, if I text her, she’ll call me right away or text me whenever she can […] say I message her past 4:30. Then she’ll get right to me right when she gets to work the next day. And that’s the best part […] I know I’m going to get an answer within the next 24 hours. (3-2-5)

When asked how they knew a nurse could be trusted, one participant highlighted the PNCC nurse’s ability to keep their conversations and work private, which was especially important in their shared small, close-knit community.

> Well, it’s a small community, as I mentioned, so there’s a tendency for people to talk. But, when I was in her office, she never talked about anybody else […] She would talk about other people in the sense of making me feel like I’m not alone in my thoughts or what I’m going through, but […] I had no idea [who it was]. I couldn’t go, “She must be talking about so-and-so.” So, it made me know that I could say anything, and she wouldn’t be spreading it around town. And that is a definite issue in a small town [Laughs.] (3-2-6)

Concerns about privacy were less of a concern in the larger, urban community, where it was less likely that the client would know other PNCC clients or interact with their nurse outside the program context.

##### Providing Holistic Care: Knowing the Community

It was important to participants that their nurse had some sort of shared connection, whether personal or communal. This was a critical factor in being open to *receiving* any support from the nurse (both socio-emotional and resource-related), and took two major forms: some clients discussed how sharing certain identities with their nurse, such as being a mother or tribal member, contributed to the development of comfort and trust, and others pointed to the nurse’s deep commitment to or embeddedness in their shared community regardless of the nurse’s own identities, which facilitated the establishment of trust.

When a PNCC nurse was a mother, it could signal that she could relate to the worries and stresses that came along with new parenthood. Sometimes, the nurse served as a role model for the client in terms of parenting experience and skills:

> Like, I really didn’t know her when I first—I just knew of her, so I really didn’t know her, but like, seeing her with my kids all the time and like, I don’t know, like building that relationship… I think that really helps like, because she’s a mom too, and she *knows*, like—she’s really good with kids herself. […] She’s a really good mom, so having like other role models like that I think really helped me because every kid needs their mom. (3-2-1)

Mothering experience was also associated with being “caring:”

> She’s caring. And that’s probably why she got into this in the first place. You know, she’s a mom and she has older kids, so—I think her youngest one still lives at home— but, you know, she’s got experience with being a mom. (1-2-6)

Several participants who had received services at the rural clinic site mentioned that the PNCC nurse being from the local community and specifically a member of the local Native American tribe contributed to their feeling supported by someone who “understood” them. One participant shared: “I think it’s easier to connect with [the nurse] because she grew up around here and she knows how life is around here […] Like, she knows what goes on around here.” (3-2-1)

Another participant detailed how the nurse’s community membership influenced her care:

> Not only is she a nurse, she’s working with social programs, and she’s also a mom. So, she just is well-rounded, [a] very well-rounded resource to have. She’s also a tribal member. We’re on the reservation here. My husband’s a tribal member. So, me moving up into this community to then… a set of information on levels that can’t even be described in a survey [laughs]. (3-2-6)

This nurse had also become quite well known throughout the community, which facilitated relationship-building and trust: “Everyone like, talks highly of her around [town]. And like— well, my sister too, she knows her too and just like, being around—[it’s] a small town and like, mostly everyone’s like, pretty much related.” (3-2-8)

As a converse to the shared identities that better enabled the development of comfort and trust, at the urban research site where the majority of PNCC nurses are white and a large proportion of clients are Black and/or Latina, one client lamented the racial discordance with her PNCC provider; this client is Black and had a white PNCC nurse. Though she had an overall positive experience in the program, she contrasted her interactions with the nurse to those with her doctor, who is also Black:

> It’s really hard to explain. I just feel less judgment, I think. And then, I don’t know, it’s just, I guess the word is just relatable, but it’s not just because she’s Black. It’s just, just—seeing someone that looks like you is just different. It’s out of the norm, honestly, and it’s pretty sad. But it’s just—it’s also like a motivation thing to me, too, because, like, “wow, she’s a Black doctor. Oh my God, that’s possible,” you know? And then it’s like, you kind of get interested in how she got to where she got and her life and experience and how she could help the community or—you know? (1-2-7)

This was the only client at the urban site who directly discussed the impact of racial (dis)concordance with her PNCC nurse, but it should be noted that most client-nurse dyads did not share a racialized identity.

### Theme 2: Connections to Needed Resources

#### a) The Meaning of Connections to Needed Resources for Clients

In addition to socio-emotional support, participants also highlighted the impact of identifying and connecting them to additional resources. These “resources” included free or reduced cost food; transportation; physical supplies such as a breast pump, diapers, basic infant clothes, and a bassinet or “pack ‘n play;” education and information; or referrals to infant development or parenting programs at the end of PNCC. A common refrain among many participants was that they were unaware of the programs and services available to them that the PNCC nurse connected them to. One participant described how her PNCC nurse was able to direct her to a plethora of resources in the community that she did not know existed.

> She had given me the resources, you know, and connected me with the right people for the smoking. I think I probably still have the folder of all the resources she had given me…I don’t know what she had asked me in the beginning. “Are you religious?” And I said, “Well, not really? But I have a religion, but I don’t…” “Well, here are some churches that, you know, could help you if you need them. But you don’t have to feel that you need to contact them.” Or, “Do you need to know what’s [available]?” There’s, you know, all the different connection that I had no idea were even [here]…I know they’re there now, but I would not have known without her. (1-2-9)

This participant further elaborated on the need for resource support because most women, like herself, are not aware of what is available to them. She also emphasized that the need for resource support is particularly true for single moms.

> I didn’t know about the different kinds of programs. I didn’t know the different phone numbers you could call. I didn’t know about the respite center. I didn’t know about all of these places, until she [PNCC nurse] had come into my life. And it was one of those things where, [chuckle] I think that’s the number one thing people need is the support because they don’t know what’s out there. And it’s, you know, I have a few friends who are single moms. They don’t have the amazing support I do from my husband, my family…I have a crib, but, you know, but I want a Pack N Play so I had asked her for a second one for down in my you know, down in my living room and “Can I have another one?” People don’t realize that that’s out there and that’s an option. (1-2-9).

In addition to getting connected to or finding out about programs and services for pregnant women, another type of resource support that many participants reported receiving was educational materials and supplies for breastfeeding:

> Like when my [baby] was in the NICU I would talk to [PNCC nurse] about it, and she would just…give me information about like breastfeeding and how I could do it from home while he was down there…it was just relieving that she had all this information that could help me when I didn’t know what else to do. (3-2-1)

The resource support provided for breastfeeding often served a two-pronged purpose of not only providing information and supplies but helping to support a participant’s goal to breastfeed:

> So, I had got a breast pump from one of my friends because I breastfeed…and it was a couple years old, and the motor stopped working in it. And she [PNCC nurse] was the first person I thought of that could help me. And within a couple of days, I had a brand-new breast pump… Like I was in need of something, and she was there right away and called who she had to call and got it set up for me. So that was very helpful. Like, that was like, the, “oh, my God,” because breastfeeding was very important to me. So, she made that so I didn’t fail at it. (1-2-6)

Even though clients deemed resource connections to be highly important, the value of this form of assistance was tied up in the emotionally supportive and trusting relationship they had with PNCC nurses. PNCC nurses were attuned to the dual needs of socio-emotional and resource support, and thus often provided them in tandem. One participant characterized her PNCC nurse as an “information box” because she was very knowledgeable about everything from pregnancy and postpartum care to community resources and material supports. However, in addition to her expertise, she also provided socio-emotional support:

> If I asked her any, like I could just talk to her about anything because she would have an answer or advice for it. Like I was having a really hard time with my husband. I talked to her about it. She had some really good advice. (3-2-1)

Another client described a triad of skills that the nurse brought to their work together; an “emotional link,” a “professional” goal setting approach, and her “contacts and resources.”

> …she [PNCC nurse] was such an emotional support for me…When I had gotten back from the hospital, I was told I cannot pick up my daughter. She was over my weight limit. And [the PNCC nurse] had showed up, and I told her, and I started bawling… I just started crying. And she’s like, “[Participant], it’s okay. You’re gonna get through it,” and, “Here. Why don’t you try some of this?” […] I mean, she was every—she stopped becoming a representative of a program, and I considered her more of a friend than anything. I mean, the emotional link of well, “I’m here. What do you need? Like, just talk to me.” The professional, “Let’s set a goal.” And the “Well here, I have contact”—you know, “my resources.” She had become an all-around “the person.” I needed a resource; I could contact her. I needed someone to cry to; I could call her […] There’s not much she did not do to help me in some way while I was pregnant. (1-2-9)

Along the same lines, another participant described the combination of resource and socio-emotional support she received:

> So, if you need help with resources in the community, she’s very helpful with that. She helps with, you know, checking the weight, making sure like the development is going properly and stuff like that, and make sure your health is good, not even just your physical health but your mental health, just like somebody to talk with and communicate with, having like a friend. Not a lot of people have that, so it was good to have someone to talk about. (1-2-6)

#### b) How Nurses Operationalized Connecting Clients to Needed Resources

Participants identified that the relative flexibility of the PNCC program and the variety of resources the PNCC nurse could connect them to made them feel that they were personally and individually supported. Participants reported a wide range of topics and needs that the PNCC nurse helped address, including breastfeeding and lactation support, planning for labor and birth, normal infant development and milestones, infant supplies such as diapers, relationship health, addiction, parenting skills, meeting basic needs for housing and food, and finding additional information about complex medical conditions or diagnoses. One client commented on this flexibility and variety when asked about the kinds of topics she discussed or issues that she worked on with her PNCC nurse:

> Just anything, everything. […] She has a Facebook that I could just message or anything. And like, every time something is wrong—I think something is wrong with my kids, I’ll message her, and she’ll give me advice on what I can do. […] There wasn’t anything that you weren’t supposed to talk about with them. (3-2-1)

The adaptability of the program to client needs was important because clients felt that the nurse was actually there to help them rather than just to deliver a program:

> She was just kinda like an outlet. […] It wasn’t more, like, a “program lady.” She was more, like, someone I can communicate to and somebody who actually, like, helps me focus on what I need to do: what’s best for my health, for my child. […] She was working hands-on, so to speak. Where, like—you know how, like, when you go to a program and they’re like, “Oh, this is the strict guidelines. Blah, blah, blah.” She was more, like, communicating with stuff. (1-2-8)

Another client contrasted the PNCC nurses’ flexibility and openness to that of the staff from other social services she had received.

> Like they don’t try to act like ICW [Indian Child Welfare] workers or social workers or anything. You know what I mean? They’re just—they’re more like, welcoming, I guess, and more approachable. I think it’s really good, this program. If it wasn’t here, I don’t know what I would have did. I can’t imagine it not here. (3-2-1)

Because of the flexibility, PNCC clients often described feeling truly listened to while interacting with their PNCC nurse.

> She was really nice and listened very well. […] I think mainly just like, telling someone how I was feeling and what I was hoping, and they were listening and even given advice as they’re asking questions… it seemed kind of comforting, I guess. (1-2-10)

Again, this was sometimes framed as different from their typical interactions with healthcare providers. The same client compared these experiences with her PNCC nurse:

> I’m just like, thinking that maybe she cared a little bit. Cause, the doctors and nurses, they’ll care, but obviously they have other patients and they’re… busy, so they can’t just sit there and listen to everything that you have to say. But I think that’s where she came in, she kinda listened to everything you had to say. (1-2-10)

### Theme 3. Client’s Recommendations for Program Improvement

#### a) Lengthen Time Frame of PNCC Program

Participants made several suggestions about ways the program could be even more supportive of them and their communities. Almost universally, clients advocated for a longer program duration. Currently, eligibility for PNCC ends when the client is 60 days postpartum, reflecting the timeline for public health insurance coverage during pregnancy. When asked about what she would change about the program, one client replied, “the only thing I would say change is be able to stay longer” (1-2-5). Participants reflected that 60 days postpartum is actually a critical time for support, with the stresses of early parenting, return to work for some clients, breastfeeding and lactation concerns, and managing the family’s health.

> I wish it was a little longer than it is. That’s what I wish. […] I wish it was a couple years longer… the only way to reconnect with them would be to get pregnant again. (3-2-4)

Most clients had been referred to additional programs and services post-PNCC participation, including Early Head Start and local parenting classes. However, these resource referrals were often less than appealing because they would require building an entirely new trusting and supportive relationship, which had been so well developed between PNCC nurse and client.

> I think the biggest thing from me, I feel, is that they should continue it, like, after your child is born. I know they kinda, like, send you to somebody else if you wanted to? But I feel we should continue with that same person if you wanted to. Because it’s like, you gotta build a whole new relationship with one person and it’s like, you may get along with one person, but not get along with the next person, you know what I mean? (1-2-8)

The length of the program and challenges with “graduating” clients before the nurses felt they were ready was also a major theme discussed in our simultaneous interviews with PNCC nurses; these findings will be discussed in a forthcoming publication.

#### b) Improve Marketing and Community Knowledge of PNCC

Although participants seemed to be satisfied with the way PNCC nurses connected them to outside resources, many clients of the urban site stressed that PNCC itself needed to be a more visible and well-known resource in their communities. Compared to the rural clinic, where almost everyone knew about the program and had personal or social network connections to the nurse before formally enrolling in PNCC, these urban-dwelling clients had not heard of PNCC before it was offered to them. Some clients had therefore been apprehensive or hesitant to enroll in PNCC.

> I had no idea this program was a thing and I think not just for me, but everybody else, I think it would’ve been more helpful just to know it existed. So like, “Can we have somebody call, call you from,” you know, “public [health]”. And I’m like, “From who? What? What are you talking about?” Like, “sure, but who are they?” And I think that was part of the reason why I wasn’t sure what to think because I didn’t know what it was. […] I mean, my doctors, I’m sure, wouldn’t give me somewhere that wasn’t reputable, but I’ve never heard of them. I didn’t know it existed. I didn’t know what it was. (1-2-9)

Like engaging with other social services and programs, this client asserted that knowing about the PNCC program beforehand may have eased some of her anxiety about enrolling.

Several participants also speculated that PNCC was not reaching all of the families that could benefit from the program and suggested that the program do more active marketing so that more community members could benefit from it.

> I honestly think, like, it should be known. Like, whether they have something at the farmers market downtown, or a billboard, or something. […] I think that there should be some literature or something in— from Planned Parenthood, all the way up to all of the hospitals. Especially for women who are high risk, especially for a single woman, and especially for a single woman with other kids, too. (1-2-8)

Other clients also suggested that PNCC could facilitate more personal and social connections and community-building among clients:

> I can’t really say nothing bad about the program. I feel like it should be more known. I just feel like more females who are younger than me, and my age, need to know about this program, because… I think it just eases people’s mind, especially females, like, whether it’s your first kid or not. But just letting you know that there’s other resources out there that can help you and letting you know that, like, there’s other people—I think they have a few, like, group things or something like that. You get together with […] like, females around the same age [to] be able to have, like, chat time—whether we’re on a video chat because of the virus or meeting up somewhere—to be like, “Hey,” like, “there’s other people that are here for you. Hey,” like, “I know what you’re going through.” I think the biggest thing that they need to do and work on is getting the word out there about this program. (1-2-8)

Building these personal and social connections might have helped clients address difficult behavioral and mental health challenges that were common in their own community:

> I think support groups, like actual, like, groups like, that help about postpartum depression and like, if you don’t know what’s wrong with your baby and it just—the baby keep crying—there’s someone there that will give you advice on what you can do and […] I just feel like, if they’re drug addicted and like, had groups, that would help them be clean—like, become clean—and wean themselves off drugs before their baby is here. Just groups like that, because there’s a lot of people that have babies that are born addicted. (3-2-1)

In addition, such peer support might have enabled an easier transition from the PNCC program through the development of more enduring friendships with others in the community. This could have prevented clients from feeling the full impact of the abrupt end of PNCC support at 60 days postpartum.

## Discussion

In contrast to the most frequently and sometimes required measured outcomes of nurse home visiting programs – i.e., physical health and birth outcomes – clients asserted that the positive mental health and overall wellbeing attained through the social and emotional support they received through PNCC engagement was of primary importance. Clients also stressed that the PNCC nurse’s ability to connect them to resources that could help meet their immediate needs was an important outcome of the program. Those connections were meaningful to many clients because they happened in the context of those socially and emotionally supportive relationships with a PNCC nurse. These findings further substantiate earlier research showing the effect and value of social support and resource connection for PNCC clients (Heitzman et al., 2019).

The socially and emotionally supportive nurse-client relationship could be generated in multiple ways, but two major mechanisms were salient in our data. First, certain nurse identities, such as being a mother and/or member of the same community, signaled to potential and new clients that a nurse was caring and/or known, and thus, could be trusted. This was true largely in our rural research site that served a smaller, closer-knit community. Second, these relationships were built over time through the nurse’s reliable presence, follow through on promised services and information, and responsiveness to client needs and circumstances. The aforementioned modes corroborate extant research pinpointing nurse characteristics and skills as key to developing a trusting nurse-client relationship (Forchuk et al., 2000; Ozaras & Abaan, 2018).

Several clients mentioned struggling with mental and behavioral health issues such as postpartum depression and addiction, and how the social support provided by the PNCC nurse helped them cope with related challenges. Mental and behavioral health outcomes have been examined in more highly structured and longer-term home visiting programs such as the Nurse Family Partnership (Beeber et al., 2021) and other MIECHV-funded programs (Tandon et al., 2020), but not in PNCC. These potential benefits or outcomes of PNCC participation are not systematically measured across PNCC programs in any state that offers this Medicaid benefit. A few PNCC programs in Wisconsin have begun to make investments in mental and behavioral health, but this has not yet been supported at the funding or policy level. Given what our findings suggest about the impact of the PNCC experience for clients, and the growing burden of mental and behavioral health concerns in the U.S., PNCC will need to move beyond traditional metrics of program effectiveness such as physical health outcomes (e.g., birth weight and preterm birth), and begin to target social, emotional, and mental health outcomes.

Our data support some recommendations that would move towards optimizing the PNCC benefit. First, PNCC sites should prioritize the recruitment of community concordant PNCC providers, whether they are nurses as in both of our research sites, or community health workers. Although we did not specifically seek to document clients’ thoughts on racial concordance with their PNCC provider (see Limitations below), the benefits of concordance with healthcare providers have been documented in other settings (Cheng, Nakash, Cruz-Gonzalez, Fillbrunn, & Alegría, 2021; Shen et al., 2018). Second, given the centrality of the nurse-client relationship, PNCC provider training should be established to further develop a nurse or community health worker’s communication, client engagement, and compassionate care abilities. Third, our data clearly demonstrate that 60 days postpartum is a non-ideal time to discontinue the services provided by PNCC, and most clients argued for a longer program. Although effective program operation is dependent on adequate funding, program duration should not be dictated by funding alone (Medicaid coverage in this case) and reflect the needs of the intended client population. Several states, including Wisconsin, where these data were collected, are currently debating extending Medicaid coverage for eligible mothers to a full year after birth. Such legislation would likely mean that the PNCC benefit could be extended to one year postpartum as well, which may contribute to positive psychosocial outcomes. Finally, PNCC programs should prioritize social support and mental and behavioral health screening and interventions. Since the flexibility and individualized approach of PNCC is so critical to its success, additional research should explore the best approaches to those improvements. PNCC providers should also systematically measure their performance on social and emotional support and related outcomes of patients.

### Limitations

Our findings should be considered in light of several limitations. First, we experienced recruitment challenges at the larger, more urban research site, with few former PNCC clients reaching out to the research team with interest in the study. The research team regularly discussed these challenges with the site’s PNCC team, discussing possible reasons for slow recruitment. After these conversations, we can guess that this difficult recruitment reflects some combination of a) some of the characteristics of the target population including frequent moves and unstable housing (thus mailed letters would not have reached them), b) contentious and damaging histories with health-related research—in this specific community and in the U.S. overall—that contributed to mistrust and unwillingness to participate, and c) our participant payment was not enough to motivate or enable the time and effort required to participate. Regardless of the explanation, the slow recruitment process may have resulted in a sample that distinct from the larger group of PNCC clients and therefore did not represent the variety of experiences that exist.

Finally, several participants were hesitant to discuss certain sensitive topics with the PI during interviews, including drug use, incarceration, and racism. Some participants alluded to these factors, either in their own or a loved one’s experience, and in the second arm of the study, the PNCC nurses identified these as some of the biggest issues in their respective served communities. This hesitancy, while understandable, likely means that some crucial PNCC experiences were not captured in our data. In future studies, participant-researcher racial concordance or community belonging among the research team may help assure participants of the privacy of their data, and specific data collection regarding these sensitive topics may be warranted.

## Conclusion

This study begins to fill in the gaps in evidence on the impact of PNCC by shedding light on outcomes and benefits from the perspective of program clients. Findings from this study highlight the centrality of promoting PNCC client’s social and emotional health through a trusting and supportive nurse-client relationship, the need for a longer program period, and the development of standards to assess program effectiveness in improving such aspects of health and related outcomes. Measuring and targeting client-oriented outcomes including social and emotional health would help capitalize on PNCC’s individualized care model and maximize its potential to improve maternal outcomes for mothers in complex, high-risk contexts.

## Data Availability

Data from the present study may be available upon reasonable request to the authors.

## Notes

### Competing Interest Statement

The authors have declared no competing interest.

### Funding Statement

This study was funded by the University of Wisconsin-Madison Institute for Clinical and Translational Research (UL1TR002373) through an Advancing Health Equity and Diversity (AHEAD) Award.

### Author Declarations

The Institutional Review Board of University of Wisconsin-Madison gave ethical approval for this work (IRB ID 2018-1151).

